# Multimodal Multiple Instance Learning for Fibroepithelial Tumor Diagnosis in Breast Ultrasound

**DOI:** 10.64898/2026.09.07.26362448

**Authors:** Farhan Fuad Abir, Mahad Ali, M Shifat Hossain, Logan R Holt, Victoria Chamberlain, Tyler Shern, Tolga Ozmen, Laura J Brattain

## Abstract

Fibroepithelial breast lesions include fibroadenomas (FAs) and phyllodes tumors (PTs), which are difficult to distinguish preoperatively due to their overlapping clinical and ultrasound features. While FAs are typically managed with observation, PTs require complete surgical excision with negative margins due to their risk of local recurrence and malignant progression. Because core needle biopsy can be inconclusive in about 15% of patients, some patients undergo unnecessary surgical procedures. Breast ultrasound (BUS) is a widely available, noninvasive imaging modality, but visual interpretation alone is limited by the overlap in lesion appearance. Artificial intelligence (AI)-based BUS offers the potential to improve the preoperative differentiation of PTs from FAs, reducing unnecessary operations while ensuring appropriate treatment for patients with PTs. Most deep learning approaches operate at the image level, whereas clinical diagnosis is made at the patient level by integrating multiple ultrasound views with clinical context. We propose a patient-level multimodal multiple instance learning (MIL) framework for fibroepithelial tumor diagnosis in breast ultrasound. Each patient is represented as a bag of BUS images encoded by an ultrasound foundation model (USFM) and aggregated using attention-based MIL to produce an image-derived patient score. Clinical variables, including age, lesion size, and echogenicity, are modeled using a gradient-boosted clinical learner. The image and clinical scores are then combined using logistic score fusion. In patient-level 5-fold cross-validation, logistic score fusion achieved 0.780 AUC-ROC for classification, improving over clinical feature-only gradient boosting (GB) by +0.033 (95% CI [+0.013, +0.053], *p* = 0.001), image-only MIL by +0.081 (95% CI [+0.042, +0.119], *p* < 0.001), and MLP-based end-to-end multimodal fusion by +0.035 (95% CI [+0.003, +0.067], *p* = 0.030). These results suggest that patient-level score fusion can combine complementary ultrasound and clinical evidence for PTs vs FAs.

## 1 Introduction

Fibroepithelial breast lesions, which include fibroadenomas (FAs) and phyllodes tumors (PTs), present a significant preoperative diagnostic challenge. While FAs are common benign lesions frequently managed with surveillance, PTs require complete surgical excision with negative margins because of their propensity for local recurrence and metastatic progression in borderline and malignant cases [1]. Consequently, when a FA cannot be confidently distinguished from a PT, surgeons often perform surgical excisions to obtain a definitive diagnosis, resulting in unnecessary surgery for many FAs. This leads to poorer cosmetic outcomes, increased healthcare costs, and higher patient morbidity [2]. Core needle biopsy is frequently inconclusive due to sampling limitations, making breast ultrasound (BUS) an attractive noninvasive imaging modality for preoperative diagnosis. Although ultrasound features, such as lesion size and echogenicity, have been associated with PTs, their substantial overlap with FAs limits the diagnostic accuracy of visual interpretation alone. Methods capable of extracting subtle imaging biomarkers from BUS have the potential to improve the preoperative differentiation of PTs from FAs. This will eventually reduce unnecessary surgical excision and improve clinical outcomes while ensuring that patients with true PTs get appropriate surgical management [3].

Deep learning has shown promise for BUS phyllodes-versus-fibroadenoma (PvF) differentiation [4–9]. However, most existing approaches operate at the individual image-level. This differs from clinical practice, where radiologists interpret multiple ultrasound views of the same lesion together with the patient’s context, such as age, lesion size, and other imaging descriptors. Multiple instance learning (MIL) is a promising choice for representing each patient as a bag of images and learning from a single patient-level label rather than treating each image independently [10–14]. MIL has also been explored in medical ultrasound applications [15–18]. However, image-only MIL does not explicitly use the structured clinical variables that physicians routinely consider. Incorporating them is also nontrivial since these are low-dimensional tabular features, a scenario where tree-based models can outperform neural networks [19].

We propose a patient-level multimodal framework that combines ultrasound image indications with structured clinical variables. To overcome the limited dataset and improve AI model generalizability, all ultrasound images are encoded by a pretrained ultrasound foundation model (USFM) [20] and aggregated with attention-based MIL into a patient-level image score, while clinical variables are modeled separately with gradient boosting (GB) to produce a clinical score. A low-capacity logistic model then fuses the two scores, preserving the strengths of each modality while enabling patient-level prediction [21]. We evaluate the framework on PvF classification. To the best of our knowledge, this is the first patient-level multiple instance learning framework for fibroepithelial tumor evaluation in breast ultrasound. Our study provides three main findings. First, patient-level aggregation performs better than image-level prediction, supporting the need to align training with clinical decision-making. Second, routine clinical variables provide a useful signal for PvF, while ultrasound images add a complementary increment when fused at the score level. Third, in this study, deeper neural networks did not provide statistically reliable gains under patient-level evaluation, motivating a simpler and more reproducible design based on the pretrained USFM model and logistic fusion.

## 2 Materials and Methods

### 2.1 Dataset Description

This retrospective study was approved by the Mass General Brigham (MGB) Institutional Review Board (Protocol 23-528). The requirement for informed consent was waived given the retrospective use of fully de-identified data. Cases were queried from the surgical pathology records of 14 hospitals and clinical sites within a single integrated healthcare network sharing common institutional infrastructure and electronic health record systems, accrued between 2003 and 2024 (PTs 2003–2024; FAs 2004–2024).

We curated 700 pathology-confirmed, fully de-identified BUS cases and excluded 12 cases with missing paired ultrasound and structured clinical data, yielding a PvF cohort of 688 (mean age 35.4 *±*12.4 years): 409 FAs and 279 PTs, with 4063 ultrasound images (2712 FA, 1351 PT). The 279 PTs included mixed subtypes (198 benign, 81 borderline/malignant), all treated as one PT class. A bag comprises all ultrasound images acquired for one patient, spanning different acquisition planes and repeated captures of the same lesion. The median bag size was 4 images (interquartile range (IQR) 2-7, range 1-56); 4 (IQR 3-8, range 1-56) for FA and 4 (IQR 2-6, range 1-23) for PT (Mann-Whitney *U, p* = 5.3 *×* 10^−4^; rank-biserial *r* = +0.154). Class medians are identical, and the difference is confined to the upper tail. Fourteen bags (2.0%) contained a single image (FA 9/409, 2.2%; PT 5/279, 1.8%).

Each case was associated with structured clinical-radiologic variables: age, preoperative ultrasound-measured lesion size, and echogenicity. All 688 had a complete set of information, and no imputation was required.

### 2.2 Multimodal MIL

As illustrated in Fig. 1, the framework first aggregates a patient’s ultrasound images into a single image-derived score, then models structured clinical variables as a separate clinical score, and finally combines the two patient-level scores. In this work, patient-level means that the model learns from all the ultrasound images acquired from one patient, with a single diagnostic label assigned to the entire set rather than to individual images.

**Fig. 1.**
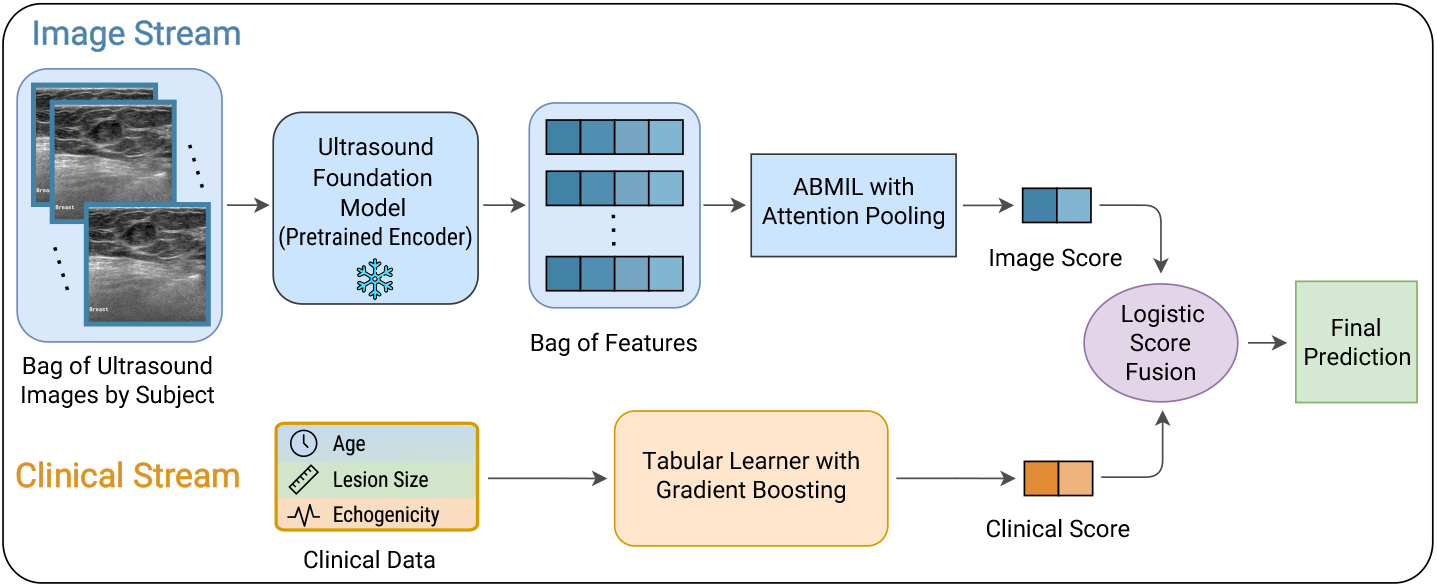
Overview of the proposed patient-level multimodal framework. A patient’s bag of ultrasound images is encoded by a pretrained USFM encoder and aggregated by attention-based MIL (ABMIL) into an image score. The structured clinical variables are modeled by a gradient boosting (GB) learner into a clinical score. A logistic score-fusion model combines the two scores into the final patient-level prediction.

#### Patient-level Ultrasound Encoding

For each patient *p*, let 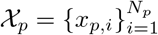 denote the bag of *N*_*p*_ BUS images belonging to that patient. In contrast to image-level classification, where each frame is treated as an independent sample, the proposed formulation assigns a single label *y*_*p*_ *∈ {*0, 1*}* to the entire bag, preserving the patient as the level of prediction. Each image *x*_*p,i*_ is passed through a pretrained ultrasound foundation model (USFM) encoder *ϕ*(*·*) [20] to get **h**_*p,i*_ = *ϕ*(*x*_*p,i*_), **h**_*p,i*_ ∈ ℝ^*d*^, *d* = 768. The encoder is kept frozen to reduce the number of trainable parameters and limit overfitting in the moderate-sized cohort. Each patient is therefore represented as a bag of fixed image embeddings, 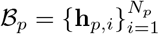.

#### Attention-based Multiple Instance Learning

Since supervision is available only at the patient level, a variable number of image embeddings must be pooled into one patient representation. We use attention-based multiple instance learning (ABMIL) [12], which learns a normalized contribution weight for each image in the bag. An attention network assigns weights *α*_*p,i*_ to the image embeddings, and the patient-level image representation is computed as 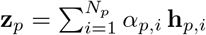. A linear prediction head maps **z**_*p*_ to an imaging score 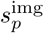, giving the imaging probability 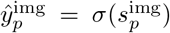. This stream is trained using patient-level labels with class-weighted binary cross-entropy to account for class imbalance. The attention weights are interpreted only as view-level contribution weights indicating which images contributed most to the patient-level prediction, and they are not interpreted as pixel-level lesion localization.

#### Structured Clinical Variable Modeling

Alongside the ultrasound images, each patient has a set of structured clinical-radiologic variables **c**_*p*_ ∈ ℝ^*m*^, including demographic/clinical context (age) and report-derived ultrasound descriptors such as preoperative lesion size and echogenicity. These variables represent routinely available clinical context used during diagnostic interpretation. Rather than fitting a neural network to this low-dimensional tabular input, we use a GB learner *g*(*·*) [22] to produce a clinical probability 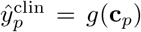. GB is well-suited to small tabular datasets because it can capture nonlinear feature interactions. Tree-based learners have also performed strongly on many tabular prediction tasks compared with neural alternatives [19].

#### Logistic Score-level Fusion

The image and clinical streams each produce a patient-level probability score, 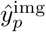 and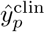. Because the clinical learner is a tree-based ensemble trained separately from the image stream, we combine the two streams at the score level using a late-fusion design [21]. A logistic model takes the two scores as input: 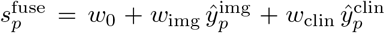, and returns the final fused probability 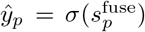. Here, logistic score fusion is used as the primary combiner to keep the final multimodal model low-capacity and interpretable. This design allows ABMIL to model variable-size ultrasound evidence, GB to model structured clinical evidence, and the logistic layer to learn a simple patient-level weighting of the two scores.

## 3 Experimental Results

### 3.1 Implementation Details

#### Image stream

Each image was resized to 224 *×*224 and normalized with ImageNet statistics. We used the pretrained USFM, a general multi-organ ultrasound foundation model, with publicly released weights and no additional in-domain pretraining [20]. In the main pipeline, the encoder is frozen, and embeddings are cached, so no encoder parameters are updated and no augmentation was used. Per patient, embeddings were aggregated by gated ABMIL (attention dim 128) [12] into an imaging score, trained with class-weighted binary cross-entropy, AdamW (lr 10^−3^, weight decay 10^−4^), and early stopping on inner-validation AUC-PR. Fine-tuning appears only in Fig. 3(a), where all encoder, pooling, and classifier parameters were updated end-to-end (AdamW, head lr 10^−3^, encoder lr 10^−4^, weight decay 10^−4^, up to 100 epochs, patience 20); the same schedule was used for all six encoders.

#### Clinical stream

Continuous clinical variables were standardized on training data. Age, lesion size, and echogenicity indicators were specified a priori as preoperative descriptors documented for all patients and supported by the prior literature. A scikit-learn GB classifier [22] produced the clinical score; learning rate *{*0.05, 0.1*}*, max depth *{*2, 3 *}*, and number of estimators *{*100, 300*}*were selected by inner-validation grid search.

#### Fusion

A logistic meta-learner combines the two stream scores into the final prediction, using scikit-learn’s default L2 penalty with *C* = 1 (no per-fold tuning). This low-capacity, two-input design keeps the multimodal model interpretable without a high-parameter end-to-end architecture.

#### Evaluation

We used a stratified, patient-grouped 5-fold cross-validation where all images from a patient remained in the same fold. Each outer fold used three disjoint, patient-grouped, class-stratified sets: inner-training (64% of the cohort), inner-validation (16%), and outer-test (20%). The ABMIL image stream and the gradient-boosted clinical stream were fitted on inner-training patients only. Inner-validation patients were used for early stopping of the image stream (AUC-PR), searching for clinical hyperparameters, fitting the logistic fusion combiner on the two stream scores, and selecting the Youden operating threshold on the fused scores. The resulting fold model was applied once to the outer-test patients, on which nothing was fitted, selected, or thresholded. AUC-ROC (primary), AUC-PR (secondary), F1, and accuracy were computed from pooled out-of-fold outer-test predictions across all five folds. The image stream used PyTorch [23]; GB and fusion used scikit-learn [24]. A fixed seed (42) was set across Python, NumPy, and PyTorch for reproducibility, and results were a single-seeded run on an NVIDIA RTX A4000 GPU.

#### Statistical testing

For each comparison, we tested the two-sided null *H*_0_: *Δ*_AUC-ROC_ = 0 (paired, same patients) with a fold-aware patient-cluster bootstrap (2000 resamples, patients stratified by class), reporting *p* as twice the smaller tail mass and 95% CIs as the 2.5/97.5 percentiles. We used a bootstrap rather than a paired *t*-test because the data are clustered by patient and AUC is a non-normal rank statistic. Here, DeLong [25] (AUC-ROC) and clustered McNemar [26] (accuracy) served as sensitivity checks. Holm correction was applied within the fusion-comparison family (*n* = 3) and the fine-tuning family (*n* = 6) [27].

### 3.2 Results

#### Ablation Studies

Table 1 reports the ablation studies on PvF classification. Image-only MIL reached an AUC-ROC of 0.699, while the clinical-only GB baseline reached 0.747. Multilayer perceptron (MLP)-based neural fusion did not improve over clinical evidence alone (0.745 vs. 0.747). The proposed logistic score fusion outperformed others, achieving 0.780 AUC-ROC and 0.725 AUC-PR. It significantly outperformed image-only MIL (*Δ* = +0.081, 95% CI [+0.042, +0.119], *p* < 0.001) and MLP-based neural fusion (*Δ* = +0.035, 95% CI [+0.003, +0.067], *p* = 0.030). Compared to the clinical structured-feature GB baseline, score fusion achieved an additional gain of *Δ* = +0.033 AUC-ROC (95% CI [+0.013, +0.053], *p* = 0.001). Overall, these results suggest that score-level fusion effectively combines complementary image-derived and structured clinical-radiologic information while remaining simpler than end-to-end neural fusion.

**Table 1.** Ablation study on PvF. Checkmarks indicate active input modalities. AUC-ROC is reported with patient-clustered bootstrap 95% CIs.

| Image | Clinical | Configuration | AUC-ROC | AUC-PR | Acc. |
| --- | --- | --- | --- | --- | --- |
| ✓ |  | Image-only MIL | 0.699 [.660,.737] | 0.622 | 0.634 |
|  | ✓ | Clinical-only GB | 0.747 [.708,.780] | 0.705 | 0.676 |
| ✓ | ✓ | Neural fusion (MLP) | 0.745 [.701,.775] | 0.708 | **0.692** |
| ✓ | ✓ | **Proposed logistic score fusion** | **0.780** [.742,.810] | **0.725** | 0.679 |

#### Patient-level aggregation

Figure 2(a) compares image-level prediction (no-MIL) with patient-level aggregation strategies. Moving from image-level prediction to ABMIL improved AUC-ROC from 0.651 to 0.699 (*Δ* = +0.048, *p* = 0.030). All MIL poolers fell within a narrow range (0.656–0.699), with ABMIL performing best. Therefore, the gain appears to come primarily from aggregating a patient’s images rather than from a specific pooling operator.

**Fig. 2.**
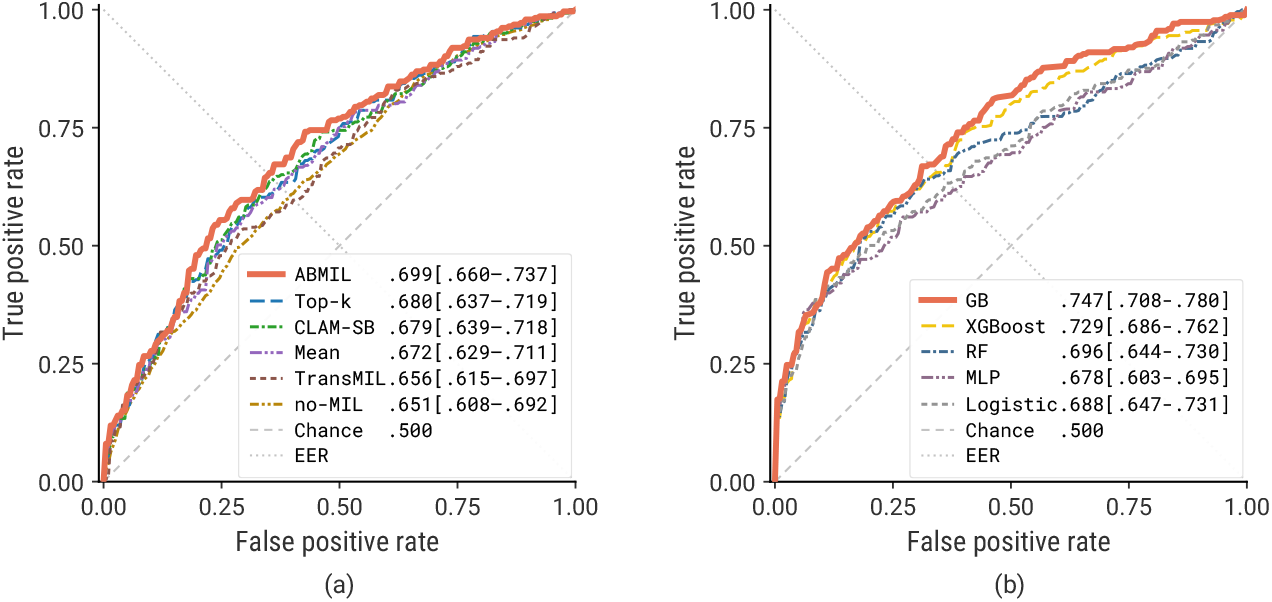
Method comparisons for the image and clinical streams on PvF classification, shown as ROC curves. Legend values report AUC-ROC with patient-clustered bootstrap 95% CIs in brackets. The equal-error-rate (EER) denotes where the false-positive rate equals the false-negative rate. (a) Image stream: image-level prediction (no-MIL) versus patient-level MIL aggregation methods, with ABMIL showing the best result. (b) Clinical stream: tree-based and boosting methods outperform linear and neural alternatives; gradient boosting was selected as the clinical learner.

#### Clinical learner comparison

Figure 2(b) compares clinical-only learners on the structured variables. Tree-based methods, namely GB (0.747), XGBoost (0.729), and random forest (RF) (0.696), outperformed logistic regression (0.688) and MLP (0.678), consistent with prior evidence that tree-based models are well suited to low-dimensional tabular data [19]. However, differences among the top boosting methods were small. Therefore, GB was selected as the clinical stream, but we do not claim that it is significantly superior to the other boosting variants.

#### Encoder choice and fine-tuning

Figure 3(a) compares six image encoders under patient-level MIL, each evaluated in pretrained and fine-tuned settings. Fine-tuning significantly degraded four of six encoders (BiomedCLIP, DINOv2, ResNet50, and ViT-B; paired patient-clustered bootstrap, Holm-corrected *p* < 0.05). For USFM, fine-tuning produced a nominal AUC-ROC increase from 0.699 to 0.716, but this improvement was not statistically significant (*Δ* = +0.017, *p* = 0.35). Moreover, Figure 3(b) compares the six pretrained encoders across AUC-ROC, AUC-PR, accuracy, and F1. USFM ranked at or near the top on all four, indicating that its selection was not an artifact of a single metric.

**Fig. 3.**
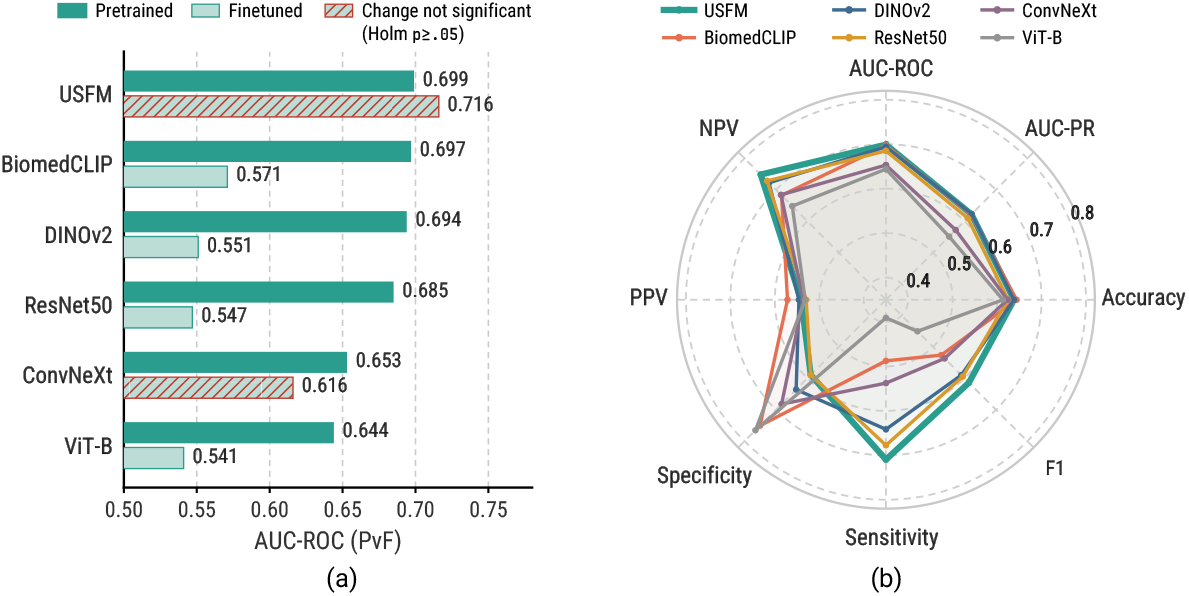
Image-encoder comparison under patient-level MIL. (a) Pretrained versus finetuned encoders; fine-tuning significantly degraded four of six encoders and gave only a non-significant nominal change for USFM. (b) Pretrained encoder performance across AUC-ROC, AUC-PR, accuracy, F1, sensitivity, specificity, PPV, and NPV, with USFM showing the most balanced performance.

## 4 Discussion and Conclusion

We proposed and evaluated a patient-level multimodal framework for distinguishing PTs from FAs using BUS images. Structured clinical variables provided a diagnostic baseline, where gradient-boosted clinical modeling achieved 0.747 AUC-ROC, above image-only MIL. Logistic score fusion achieved 0.780 AUC-ROC, outperforming clinical-only, image-only, and MLP-based multimodal fusion. Since FAs and PTs overlap in clinical profile and sonographic appearance, improvement by the image modality can help re-rank borderline cases near the decision threshold. The model is thus best viewed as combining a clinical prior with complementary ultrasound evidence rather than replacing the clinical context with image-only prediction.

Another important finding was that simpler models were better suited to this modest-sized cohort. Patient-level aggregation improved over image-level prediction, but different MIL pooling operators performed similarly, indicating that the benefit came from the patient-level problem formulation, not a specific pooling design. More elaborate aggregators, such as TransMIL and CLAM, did not improve upon attention pooling since they are designed for sparse signals across thousands of whole-slide patches. Their added capacity offers limited benefit here and may instead increase overfitting when each patient contributes only a few ultrasound views. Similar trends were observed beyond MIL. GB outperformed linear and neural models on the small clinical feature set, and finetuning large encoders gave no statistically reliable gains. Although fine-tuning can adapt pretrained encoders to the target task, it likely increased overfitting in this modest-sized patient-level cohort. Using a pretrained USFM encoder allowed the image stream to leverage ultrasound-specific modality context learned from larger-scale ultrasound data, which is especially useful in our modest-sized PvF cohort. Overall, these findings support a simpler pipeline based on pretrained USFM embeddings, patient-level MIL, gradient-boosted clinical modeling, and logistic score fusion. Bag size was also a potential confounder because FA cases had more images than PT cases. However, if we use bag size as a feature for PvF classification, the AUC-ROC is only 0.577 (95% CI [0.535, 0.619]), well below the image stream AUC-ROC of 0.699. Moreover, ABMIL attention weights sum to one, so the pooled representation is not influenced by the bag size.

However, several limitations remain. The cohort is retrospective and drawn from 14 sites within one healthcare network; therefore, acquisition heterogeneity by BUS operator or hospital site is confined to a single health system and region. The two-decade accrual window also spans several generations of ultrasound equipment, which contributes to high variability of BUS image quality. External validation on an independent cohort remains necessary. The current set of clinical variables is also limited. Additional information, such as core needle biopsy pathology, should be considered. Finally, the framework used USFM as the only ultrasound-specific pretrained encoder. Alternative ultrasound-specific foundation models should also be evaluated.

In summary, our patient-level multimodal framework combined a clinical baseline with MIL through score-level fusion, favoring a simple and interpretable strategy over complex architectures. Future work will expand the framework to include larger datasets, incorporate uncertainty estimation, and combine additional modalities such as mammograms and histopathology.

## Data Availability

All data produced in the present study are available upon reasonable request to the authors

